# Persisting influence of structured early childhood education exposure on pre-adolescent cognition – Evidence from an Indian birth cohort

**DOI:** 10.1101/2025.10.20.25338401

**Authors:** Beena Koshy, Manikandan Srinivasan, Rachel Beulah, Nandhini Mani, Venkat Raghav Mohan, Sushil John, Gagandeep Kang

## Abstract

**Background and objectives:** Structured early childhood education (ECE) can enhance human potential by improving academic performance, particularly in impoverished settings. This study evaluated the association between ECE and cognition at age 12 years in the longitudinal malnutrition and enteric diseases (MAL-ED) cohort follow-up study in Vellore, South India.

**Methods:** The MAL-ED Vellore cohort enrolled 251 newborns between 2010–2012 from urban low- and middle-income (LMIC) settings and followed them at multiple timepoints. Parents reported ECE details, including duration of enrollment. At 12 years, cognition was assessed using Malin’s Intelligence Scale for Indian Children (MISIC). Bivariate and multivariate regression analyses were conducted with adjustments for socioeconomic position (SEP), sex, maternal education, and stunting status.

**Results:** Of the original cohort, 212 (84.46%), 205 (81.67%), and 191 (76.09%) children were available at 5, 9, and 12 years, respectively. By age 5 years, 54.25% (115/212) had attended structured ECE. Exposure for 18–24 months was positively associated with verbal IQ [β: 4.36 (1.85–6.86)], performance IQ [β: 4.96 (1.25–8.68)], and total IQ [β: 4.66 (1.96–7.36)] at age 12 years, compared to those without exposure, after adjustment for SEP, sex, stunting at 2 and 5 years, and maternal cognition.

**Conclusions:** Twelve-year follow-up of an Indian birth cohort demonstrated that structured ECE continued to show positive associations with pre-adolescent cognition, extending evidence previously reported at 5 and 9 years. Structured ECE and its effective implementation may optimize human capital in India.

## Introduction

Cognition, or intelligence, has been defined as a “combination of multiple abilities in a child” by Howard Gardner [1]. The first 1,000 days of life, beginning with the antenatal period, continuing through the perinatal period, and extending into the first 2 years, are crucial for cognitive development, as most brain growth and maturation occur during this time [2]. Factors such as early childhood malnutrition, infections, perinatal asphyxia, iron deficiency, lead toxicity, and poverty adversely affect cognitive development in children [3–5]. Among these, poverty is the most significant contributor, as children from low-income households are more likely to experience unsupportive environments and limited caregiver engagement.

Early childhood is a sensitive developmental period characterized by rapid brain growth through neural network formation, myelination, and pruning. This process underscores the importance of both the “First 1,000 days” (conception–2 years) and the “Second 1,000 days” (2–5 years) [1]. Evaluating early childhood development requires attention not only to interconnected influences such as nutrition, nurturing care, and supportive environments, but also to the timing and sequences of these influences to plan for effective interventions. The Nurturing Care Framework (NCF) proposed by the World Health Organization (WHO)— encompassing adequate nutrition, health, safety and security, responsive caregiving, and early learning opportunities—provides a foundation for strengthening early childhood development [2].

In low- and middle-income countries (LMICs), childhood malnutrition contributes to morbidity, mortality, and stunting, thereby diminishing human potential [3]. The Malnutrition and Enteric Diseases (MAL-ED) India cohort, which forms the basis of this analysis, reported declines in developmental scores between 6 and 36 months across cognition, language, motor, and social domains [4]. Associated factors included low socioeconomic position (SEP), undernutrition, stunting, iron deficiency, maternal cognition, maternal mental health, and home environmental influences such as caregiver responsivity and access to appropriate play materials. Other studies have highlighted similar factors—perinatal asphyxia, infections, micronutrient deficiencies, heavy metal toxicity, maternal depression, and poverty—as detrimental to human potential, with poverty exerting the greatest influence by intersecting with many of these risks [5–7].

In LMICs, structured early childhood education (ECE) can mitigate these adverse influences and enhance developmental outcome [8]. Quality ECE has been shown to improve not only educational and economic trajectories but also health outcomes, adult morbidity patterns, crime rates, and quality of life, thereby contributing to overall “life-cycle health” [9,10]. In disadvantaged LMIC environments, high-quality birth-to-5-year ECE programs yield a 13.7% annual return, with a benefit–cost ratio of 7.3 [11]. Within the MAL-ED India study, structured ECE during preschool years positively influenced verbal, performance, processing speed, and overall cognitive abilities at 5 years, with predictive associations for verbal, performance, and full-scale cognition at 9 years, independent of SEP, maternal cognition, stunting, and home environment factors. Comparable findings have been reported in Bangladesh and Brazil [12– 14]. While some early childhood influences may diminish as children age and encounter new challenges and opportunities, evidence suggests sustained benefits. With this background, the current analysis evaluated associations between structured ECE in the MAL-ED India cohort and their cognitive abilities in the preadolescent age group of 12 years. It was hypothesized that ECE would continue to have an influence in this age group.

## Methods

This analysis was conducted in the MAL-ED India cohort follow-up, originally recruited as part of a multinational study [15]. The Vellore site in India contained densely populated urban slums, the details of which have been published previously [16–19]. In summary, 251 newborns were enrolled with parental consent between 2010–2012 and followed up at multiple time points [17]. Recruitment and follow-up collected extensive data, including sociodemographic information, childhood morbidity patterns, anthropometry, and developmental measures. The Ethics Review Committee and Institutional Review Board of Christian Medical College, Vellore, approved both the initial recruitment and subsequent follow-up studies.

### Measures

#### 1. Anthropometry

Trained field workers measured infant length using an infantometer until age 2 years and height using a stadiometer until age 9 years. Height z-scores were calculated with the Multicentre Growth Reference Study (MGRS) standards until age 5 years and with WHO AnthroPlus software for the 9-year assessment [20]. Stunting was defined as HAZ < –2 on WHO growth curves [20], and catch-up growth was defined as HAZ within normal limits after prior stunting [21].

#### 2. The SEP (Socioeconomic position)

The SEP was assessed by trained field workers using the Water and sanitation, Assets, Maternal education, and total household Income (WAMI) composite measure [22].

#### 3. Maternal cognition

A trained psychologist assessed maternal cognition at 6–8 months of the child’s age using Raven’s Progressive Matrices [23]. Raw scores were used in analyses.

#### 4. Structured ECE

At age 5 years, caregivers provided information on the number of months of structured ECE received in Anganwadi or kindergarten settings [12]. When possible, this information was corroborated with family admission records.

#### 5. Pre-adolescent cognition

At age 12 years, a trained psychologist assessed cognition at a community study clinic using Malin’s Intelligence Scale for Indian Children (MISIC) [24]. The MISIC, adapted from the Wechsler intelligence scales, included verbal and performance domains, yielding verbal (VIQ), performance (PIQ), and full-scale (FSIQ) scores.

### Statistical analysis

Data collected by field research assistants were validated by supervisors and entered in the central MAL-ED database. Categorical variables such as child’s sex, SEP, and stunting status at ages 2 and 5 years were presented as percentages. Children with WAMI scores ≥33rd percentile were categorized as high SEP. Based on stunting status at ages 2 and 5 years, children were classified as never stunted, stunted at 2 years with catch-up by 5 years, or stunted at both ages. The exposure of interest, ECE attendance, was categorized by tertiles: group 1 with no ECE attendance, group 2 with 1–17 months, and group 3 with 18–24 months. The outcome, domain-specific IQ scores at 12 years, was visualized with box plots by ECE group. After assessing normality of IQ outcomes with the Shapiro–Wilk test, bivariate and multivariable linear regression analyses were performed to examine associations with ECE attendance. Model fit was evaluated using R^2^ values. Beta coefficients and 95% confidence intervals (CI) were reported, and statistical significance was set at p < 0.05. Analyses were conducted with STATA version 16 (Stata Statistical Software: Release 14, StataCorp LP, College Station, TX).

## Results

This birth cohort enrolled 251 newborns as part of the MAL-ED study at the Vellore site during 2010–2012, as described by Koshy, et al. [18]. Sociodemographic variables, including sex and socioeconomic position, did not differ significantly between baseline and follow-up assessments at ages 2 and 5 years [18]. Cohort follow-up rates at ages 5, 9, and 12 years were 212 (84.46%), 205 (81.67%), and 191 (76.09%), respectively, with migration out of the study area being the most frequent reason for dropout. The 9-year assessments were conducted during 2019–2021, and the 12-year assessments during 2022–2024. The sex distribution of study children remained comparable across assessments at 0, 5, 9, and 12 years (P > 0.05). Stunting analysis showed that children were distributed as follows: never stunted, 113 (53.30%); stunted at 2 years but caught up by 5 years, 41 (19.34%); and stunted at both 2 and 5 years, 58 (27.36%) (data not shown).

By age 5 years, 54.25% (115/212) of children had received structured ECE, with a cohort median (IQR) of 8 (0–18) months. Classification of ECE attendance by tertile cutoffs yielded group 1 (no ECE), 97 (45.75%); group 2 (1–17 months), 55 (25.94%); and group 3 (18–24 months), 60 (28.30%). At age 12 years, mean (SD) IQ scores were 86.78 (7.22) for verbal, 92.50 (10.59) for performance, and 89.64 (7.91) for total IQ. Stratified analysis indicated that children in group 3 had higher verbal IQ [mean (SD): 90.21 (6.60)] compared with group 1 [85.39 (7.21)]. Similarly, performance IQ was higher in group 3 [97.10 (6.57)] than in group 1 [90.27 (11.17)] (Fig 1).

**Fig 1.**
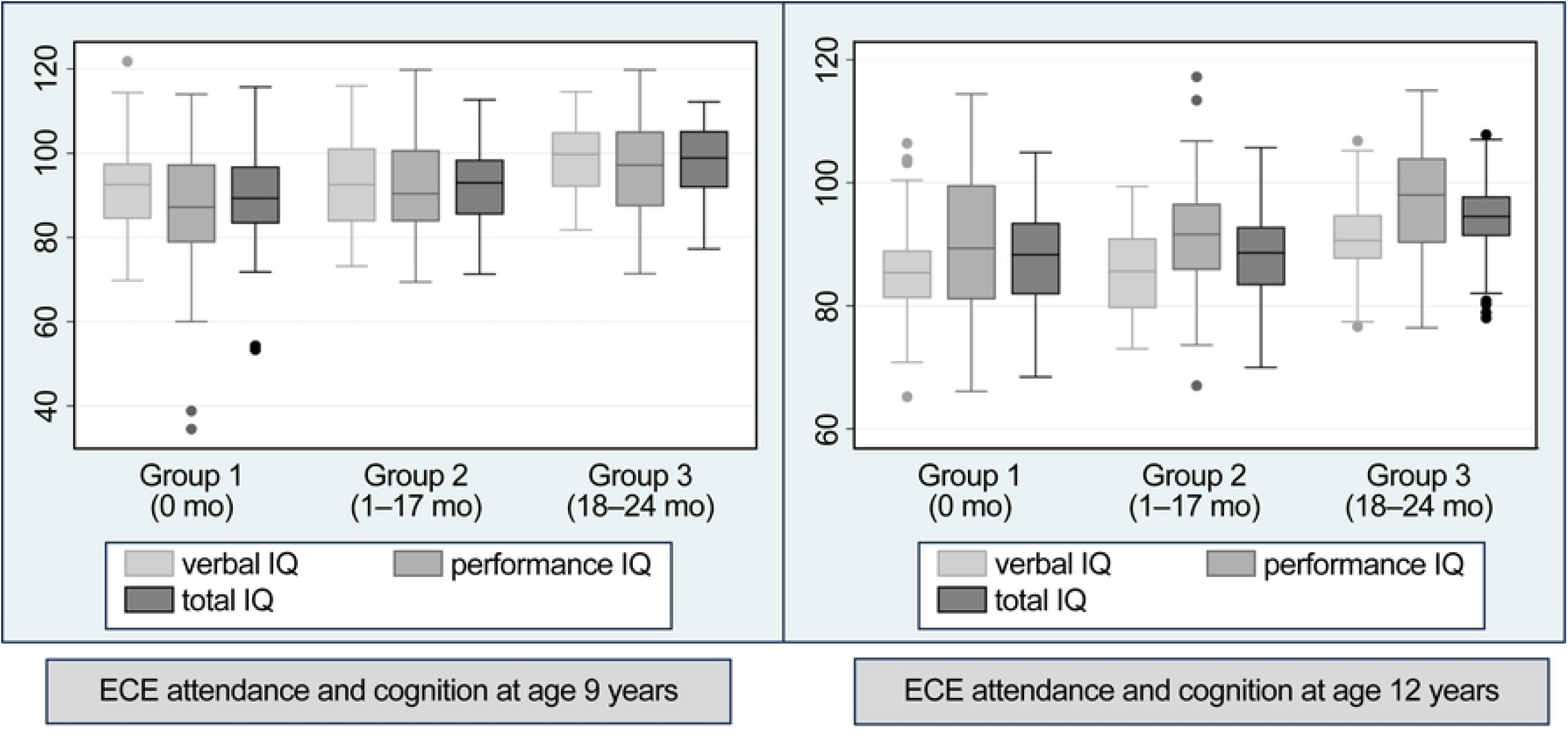
Group-wise comparison based on ECE attendance and cognition in children from the MAL-ED cohort at ages 9 and 12 years.

Structured ECE attendance was positively associated with verbal, performance, and total IQ scores at age 12 years. Unadjusted analysis showed that children with 18–24 months of ECE had higher IQ scores, particularly in the performance domain, compared with those with no ECE attendance (Table 1). Multivariable analysis further demonstrated that children with 18– 24 months of ECE had significantly higher IQ scores than those with no ECE, after adjustment for maternal cognition, SEP, and stunting at ages 2 and 5 years. Specifically, structured ECE of 18–24 months was associated with higher verbal IQ [β: 4.36 (1.85–6.86)], performance IQ [β: 4.96 (1.25–8.68)], and total IQ [β: 4.66 (1.96–7.36)] compared with no ECE attendance (Table 2).

**Table 1.**
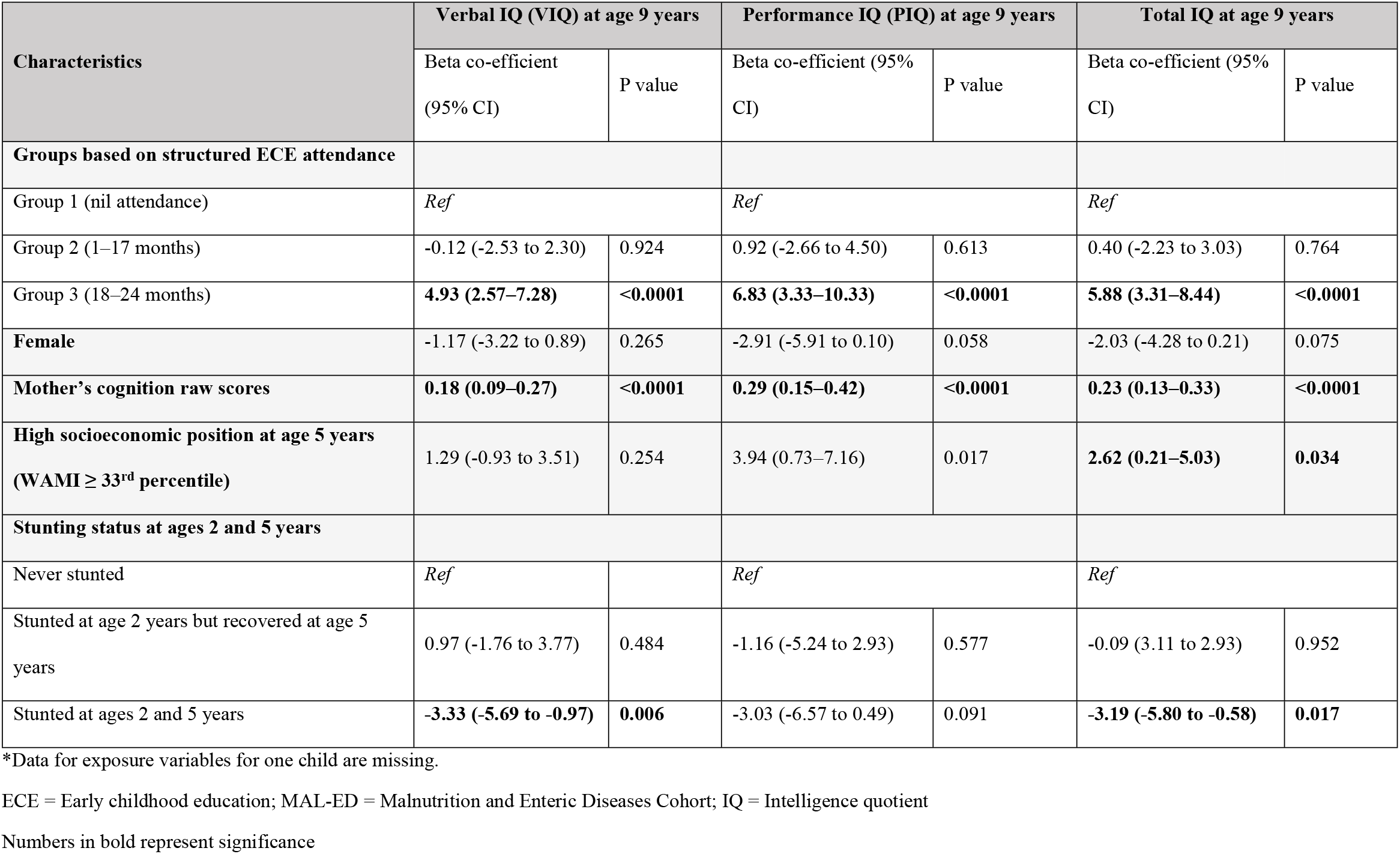
Bivariate analysis of the association between structured ECE attendance and cognition at age 12 years among children in the MAL-ED cohort (N = 190*)

| Characteristics | Verbal IQ (VIQ) at age 9 years |  | Performance IQ (PIQ) at age 9 years |  | Total IQ at age 9 years |  |
| --- | --- | --- | --- | --- | --- | --- |
|  | Beta co-efficient<br>(95% CI) | P value | Beta co-efficient (95%<br>CI) | P value | Beta co-efficient (95%<br>CI) | P value |
| **Groups based on structured ECE attendance** |  |  |  |  |  |  |
| Group 1 (nil attendance) | *Ref* |  | *Ref* |  | *Ref* |  |
| Group 2 (1–17 months) | -0.12 (-2.53 to 2.30) | 0.924 | 0.92 (-2.66 to 4.50) | 0.613 | 0.40 (-2.23 to 3.03) | 0.764 |
| Group 3 (18–24 months) | **4.93 (2.57–7.28)** | **<0.0001** | **6.83 (3.33–10.33)** | **<0.0001** | **5.88 (3.31–8.44)** | **<0.0001** |
| **Female** | -1.17 (-3.22 to 0.89) | 0.265 | -2.91 (-5.91 to 0.10) | 0.058 | -2.03 (-4.28 to 0.21) | 0.075 |
| **Mother's cognition raw scores** | **0.18 (0.09–0.27)** | **<0.0001** | **0.29 (0.15–0.42)** | **<0.0001** | **0.23 (0.13–0.33)** | **<0.0001** |
| **High socioeconomic position at age 5 years**<br>**(WAMI ≥ 33<sup>rd</sup> percentile)** | 1.29 (-0.93 to 3.51) | 0.254 | 3.94 (0.73–7.16) | 0.017 | **2.62 (0.21–5.03)** | **0.034** |
| **Stunting status at ages 2 and 5 years** |  |  |  |  |  |  |
| Never stunted | *Ref* |  | *Ref* |  | *Ref* |  |
| Stunted at age 2 years but recovered at age 5<br>years | 0.97 (-1.76 to 3.77) | 0.484 | -1.16 (-5.24 to 2.93) | 0.577 | -0.09 (3.11 to 2.93) | 0.952 |
| Stunted at ages 2 and 5 years | **-3.33 (-5.69 to -0.97)** | **0.006** | -3.03 (-6.57 to 0.49) | 0.091 | **-3.19 (-5.80 to -0.58)** | **0.017** |

\*Data for exposure variables for one child are missing.
ECE = Early childhood education; MAL-ED = Malnutrition and Enteric Diseases Cohort; IQ = Intelligence quotient
Numbers in bold represent significance

**Table 2.** Multivariate analysis measuring association between structured ECE attendance and cognition at age 12 years among children of the MAL-ED cohort (N = 190*)

| Characteristics | Verbal IQ (VIQ) at 9 years |  | Performance IQ (PIQ) at 9 years |  | Total IQ at 9 years |  |
| --- | --- | --- | --- | --- | --- | --- |
|  | Beta co-efficient (95% CI) | P value | Beta co-efficient (95% CI) | P value | Beta co-efficient (95% CI) | P value |
| **Groups based on structured ECE attendance** |  |  |  |  |  |  |
| Group 1 (nil attendance) | *Ref* |  | *Ref* |  | *Ref* |  |
| Group 2 (1–17 months) | -0.46 (-2.80 to 1.88) | 0.174 | -0.14 (-3.61 to 3.33) | 0.936 | -0.30 (-2.82 to 2.22) | 0.814 |
| Group 3 (18–24 months) | **4.36 (1.85–6.86)** | **0.001** | **4.96 (1.25–8.68)** | **0.009** | **4.66 (1.96–7.36)** | **0.001** |
| **Female** | -1.37 (-3.34 to 0.61) | 0.174 | **-3.46 (-6.38 - -0.54)** | **0.021** | **-2.41 (-4.53 to -0.29)** | **0.026** |
| **Mother's cognition raw scores** | **0.18 (0.08–0.27)** | **<0.0001** | **0.28 (0.15–0.42)** | **<0.0001** | **0.23 (0.13–0.33)** | **<0.0001** |
| **High socioeconomic position at 5 years (WAMI ≥ 33<sup>rd</sup> percentile)** | -1.61 (-3.86 to 0.67) | 0.165 | 0.80 (-2.56 to 4.16) | 0.639 | -0.40 (-2.84 to 2.04) | 0.745 |
| **Stunting status at ages 2 and 5 years** |  |  |  |  |  |  |
| Never stunted | *Ref* |  | *Ref* |  | *Ref* |  |
| Stunted at age 2 years but recovered at age 5 years | 0.62 (-1.95 to 3.20) | 0.633 | -2.08 (-5.89 to 1.73) | 0.284 | -0.73 (-3.50 to 2.05) | 0.606 |
| Stunted at ages 2 and 5 years | **-2.55 (-4.87 to -0.23)** | **0.031** | -1.85 (-5.28 to 1.59) | 0.291 | -2.20 (-4.69 to 0.30) | 0.084 |

\*Data for exposure variables for one child are missing.
R2 values for regression models for verbal IQ, performance IQ, and total IQ include 16.59%, 14.97%, and 19.53, respectively.
ECE = Early childhood education; MAL-ED = Malnutrition and Enteric Diseases Cohort; IQ = Intelligence quotient
Numbers in bold represent significance

## Discussion

This prospective follow-up study among children living in an urban Indian LMIC setting evaluated associations between structured ECE and pre-adolescent cognition at age 12 years. A prior analysis of the same cohort reported effect sizes ranging from 3.32 to 19.55 for concurrent associations between ECE and cognition at age 5 years, and from 5.25 to 8.82 for predictive associations with cognition at age 9 years, even after adjustment for SEP, stunting status, maternal cognition, and home environment [12]. The present study found effect sizes of 4.36, 4.96, and 4.66 for VIQ, PIQ, and FSIQ, respectively, in a similarly adjusted model.

Early childhood poverty and related adversities hinder an optimal learning environment both at home and in preschool. The MAL-ED India cohort demonstrated a decline across developmental domains from 6 to 36 months, influenced by SEP, stunting, iron status, maternal depression, and home environmental factors, consistent with published literature [4–7,25]. Globally, more than 200 million young children, primarily living in LMICs, experience reduced potential due to poverty and early stunting [3,25,26].

These detrimental impacts can be mitigated through early childhood support within the NCF, including adequate nutrition, safe and secure environments, good health, responsive caregiving, and opportunities for early learning. Such support helps counter poverty and environmental adversities, as neuroplasticity enables the developing brain to reorganize through new circuits and connections. ECE, which provides opportunities for early learning, contributes to improved “life-cycle health” through better well-being and quality of life [8–10]. The Heckman curve highlights the benefits of ECE investment, estimating an annual return rate of 13% [11,27].

Evidence from LMICs demonstrates the benefits of structured ECE in disadvantaged settings. In Brazil, children attending nursery schools scored 8 points higher in follow-up assessments of cognitive ability [13]. A study in Bangladesh found that preschool attendance improved primary school readiness [14]. Similarly, research in Uruguay showed both concurrent and predictive cognitive benefits at school entry and 6 years later [28]. Long-term follow-up studies, such as the 1991 NICHD study of Early Childcare and Youth Development and the Abecedarian low-SEP cohort, indicate that structured ECE can have enduring effects into adulthood, including improved education, employment, and vocational outcomes [29,30].

Despite these findings, some evidence suggests possible fadeout of ECE benefits in later childhood and adulthood [8]. Criticism of the Heckman equation includes its narrow focus on economic potential and cognitive outcomes, limited definitions of success, and inadequate attention to social, community, and family influences, as well as root causes of inequality. For example, the Tennessee Prekindergarten Program, a randomized controlled trial (RCT), demonstrated positive effects at age 4 years, but these diminished by grade 3, with some negative associations including higher rates of disciplinary issues and special education placement [31]. Similarly, the Head Start Program, another RCT, found positive effects of ECE at age 4 years, followed by fadeout by grade 3 [32]. However, subgroup analyses revealed that high-risk groups experienced better developmental and cognitive outcomes [33].

In this context, the present study demonstrates persisting effects of structured ECE on pre-adolescent cognition in an Indian LMIC birth-cohort follow-up. Compared with effect sizes at ages 5 and 9 years, the influence at 12 years was smaller; however, a significant association remained after adjustment for SEP, maternal factors, and stunting patterns. The “cognitive advantage hypothesis,” wherein children from disadvantaged backgrounds capitalize on improved environments, may underlie these outcomes in this LMIC urban slum setting [12,29].

In India, the National Education Policy (NEP) 2020 emphasizes ECE within the Integrated Child Development Services (ICDS) program, in line with UNICEF recommendations [34]. Under the “Poshan 2.0” and “Mission Saksham Anganwadi” programs, improved nutrition and ECE are being implemented in 1.3 million Anganwadi centers nationwide. In 2023, the Ministry of Women and Child Development (WCD) launched “Poshan Bhi Padhai Bhi” (PBPB); education along with nutrition, introducing two curricula for early years: “Navchetana” for ages 0–3 years and “Aadarshila” for ages 3–6 years. Effectiveness of ECE delivery can be enhanced by structured training and refresher programs for Anganwadi workers (AWWs) and by introducing support staff, as shown in studies from Maharashtra and Tamil Nadu [35,36]. In Maharashtra, initial and refresher trainings for AWWs, combined with financial support for toys and play materials, led to a 10-point increase in IQ scores [36]. In Tamil Nadu, the addition of a dedicated ECE worker improved growth parameters and cognitive performance in language, mathematics, and executive function 18-months post-intervention [35]. Elements of these support systems, including AWW training and play-based approaches, are incorporated into the PBPB program under the Ministry of WCD.

Limitations of the present analysis include the relatively small sample size within a localized population in South India and the absence of detailed information regarding ECE quality. Furthermore, the cognitive assessments emphasized logical and mathematical intelligence, without evaluating emotional or social domains that are critical in a life-course perspective. Strengths of the study include robust follow-up, standardized assessments, and the use of India-specific cognitive measures.

In conclusion, this analysis demonstrates the persisting effect of structured ECE on pre-adolescent cognition in resource-constrained settings in India, underscoring the importance of early learning support during this critical developmental window. These findings align with existing literature and extend evidence from the same birth cohort regarding associations between ECE and cognition at ages 5 and 9 years. Collectively, this evidence strengthens the rationale for integrating the Nurturing Care Framework into early childhood initiatives to maximize human capital in India.

## Data Availability

All MAL-ED data till 5 years is available at https://clinepidb.org/ce/app. The anonymized version of the MAL-ED Data including MRI Brain parcellation data is available in the Harvard Dataverse database. Below is the link to our data: https://doi.org/10.7910/DVN/KJGMAY Anonymized dataset for this analysis is also uploaded.

https://doi.org/10.7910/DVN/KJGMAY

## Acknowledgements

The authors thank the children, their families, and the staff of the MAL-ED Network Project. We would like to thank Editage [http://www.editage.com] for editing and reviewing this manuscript for English language.

## Author contributions

Beena Koshy, Manikandan Srinivasan, Venkata Raghava Mohan, and Gagandeep Kang contributed to study conception and design. Material preparation, data collection, and analysis were conducted by Beena Koshy, Manikandan Srinivasan, Rachel Beulah, Nandhini, Venkata Raghava Mohan, and Sushil John. All authors read and approved the final manuscript.

## Ethics approval

This study was approved by the Institutional Review Board of Christian Medical College, Vellore, for both initial recruitment and follow-up visits (IRB 6769 for the original cohort and follow-up; IRB 11821 for the 9-year follow-up; IRB 14380 for the 12-year follow-up). Participants were enrolled after written informed consent and assent were obtained. This research was conducted in accordance with the World Medical Association Declaration of Helsinki.

